# ARTIFICIAL INTELLIGENCE TOOLS FOR EFFECTIVE MONITORING OF POPULATION AT DISTANCE DURING COVID-19 PANDEMIC. RESULTS FROM AN ITALIAN PILOT FEASIBILITY STUDY (RICOVAI-19 STUDY)

**DOI:** 10.1101/2022.02.04.22270087

**Authors:** Marco Mazzanti, Aldo Salvi, Stefania Giacomini, Elisabetta Perazzini, Cinzia Nitti, Susanna Contucci, Massimo D’Angelo, Danilo Ballanti, Domenico Ursino, Matteo Marcosignori, Raniero Romagnoli, Marcello Tavio, Andrea Giacometti, Serena Tomassetti, Riccardo Bonazzi, Matteo Maccioni, Lina Zuccatosta, RICOVAI-19 Study

## Abstract

In order to reduce the burden on healthcare systems and in particular to support an appropriate way to the Emergency Department (ED) access, home tele-monitoring patients was strongly recommended during the COVID-19 pandemic. Furthermore, paper from numerous groups has shown the potential of using data from wearable devices to characterize each individual’s unique baseline, identify deviations from that baseline suggestive of a viral infection, and to aggregate that data to better inform population surveillance trends. However, no evidence about usage of Artificial Intelligence (AI) applicatives on digitally data collected from patients and doctors exists. With a growing global population of connected wearable users, this could potentially help to improve the earlier diagnosis and management of infectious individuals and improving timeliness and precision of tracking infectious disease outbreaks.

During the study RICOVAI-19 (**RICOV**ero ospedaliero con strumenti di **A**rtificial Intelligence nei pazienti con **COV**id-19) performed in the Marche Region, Italy, we evaluated 129 subjects monitored at home in a six-months period between March 22, 2021 and October 22, 2021. During the monitoring, personal on demand health technologies were used to collect clinical and vital data in order to feed the database and the machine learning engine. The AI output resulted in a clinical stability index (CSI) which enables the system to deliver suggestions to the population and doctors about how intervene.

Results showed the beneficial influence of CSI for predicting clinical classes of subjects and identifying who of them need to be admitted at ED. The same pattern of results was confirming the alert included in the decision support system in order to request further testing or clinical information in some cases.

In conclusion, our study does support a high impact of AI tools on COVID-19 outcomes to fight this pandemic by driving new approaches to public awareness.

## 1. Introduction

The name “coronavirus” was first mentioned in the scientific world in the year 1968, and by 1975, it was recognized by the International Committee on the Taxonomy of Viruses (ICTV).^1^ Coronaviruses causes different diseases that infect different animals and humans over the years, thereby making humans and animals as reservoir species. Importantly, coronaviruses diseases can be characterized by of respiratory failure in complicated cases of infection in humans, while central nervous system and gastrointestinal problems might arise in severe cases as a result of neural tropism, and presence in the feces has also been shown in certain cases.^2,3^

Furthermore, current evidence suggests that with many factors, atypical symptoms such as delirium, falls, generalized weakness, malaise, functional decline, and conjunctivitis, anorexia, increased sputum production, dizziness, headache, rhinorrhea, chest pain, hemoptysis, diarrhea, nausea, vomiting, abdominal pain, nasal congestion, and anosmia, tachypnea, unexplained tachycardia, or hypotension may be the suggestive presenting clinical presentation of coronavirus disease 2019 (COVID-19) in older adults.^4–6^

The unprecedented COVID-19 pandemic is caused by infection with a viral pathogen, the nascent severe acute respiratory syndrome coronavirus 2 (SARS-CoV-2) and is characterized by respiratory failure in severe cases. The communicability period of COVID-19 varies, but it is usually between 2-14 days, and the major signs and symptoms of COVID-19 are high temperature (fever), severe and consistent cough, and serious breathing problems in severe cases, however, complications result in the serious inflammation of the lungs, and organ malfunction occurs in patients with comorbidities and health conditions especially diabetes, heart disease and obesity. Like other RNA viruses, SARS-CoV-2, while adapting to their new human hosts, is prone to genetic evolution with the development of mutations over time, resulting in mutant variants that may have different characteristics than its ancestral strains. Several variants of SARS-CoV-2 have been described during the course of this pandemic, among which only a few are considered variants of concern (VOCs) by the WHO, given their impact on global public health. Based on the recent epidemiological update by the WHO, as of December 11, 2021, five SARS-CoV-2 VOCs have been identified since the beginning of the pandemic: Alpha (B.1.1.7): first variant of concern described in the United Kingdom (UK) in late December 2020; Beta (B.1.351): first reported in South Africa in December 2020; Gamma(P.1): first reported in Brazil in early January 2021; Delta (B.1.617.2): first reported in India in December 2020; Omicron (B.1.1.529): first reported in South Africa in November 2021 With high virulence and contagious nature of SARS-CoV-2, over 359 million confirmed cases and over 5 million deaths have occurred as a result of COVID-19 globally, with over 278 million recovered/discharged as of 26^th^ January 2021 ^7^. The severe symptoms associated with SARS-CoV-2 infections and the critical course of the disease have resulted in severe pressure from patients on emergency rooms and hospital facilities.

### Evidence before this study

During the last three-to-five years, digital information around the world has more than doubled and this trend is set to increase, with an exponential phenomenon that generates huge amounts of electronic clinical and imaging data: Big Data^8, 9^. Medicine is one of the main protagonists of this growth: the Big Data of health are increasing at a higher percentage than other sectors. One of the important phenomena is represented by the explosion of the IOT (Internet Of Things) which, in general, includes all the objects of common use that in favour of the technological evolution become smart and incorporating intelligent sensors capable to collect a great variety of information and transmit it to the internet network ^10,11^. In medicine, this trend refers to sensors that detect information from the human body in real time depicting an area that is assuming such importance and specificity to be leballed with a own name: IoMT (Internet of Medical Things) ^12–14^. We are already using smartwatches and wrist bands detecting data such as heart rate, temperature and movements, but now a revolution is underway which will pervasively produce wearable, implantable instruments capable of capturing information about physical, mechanical, chemical and electromagnetic feature^15–19^.

One of the advantages of IoMTs is the possibility of promoting adherence to hospitalizations and therapies, given that tools for sharing data between doctor and patient have been a reality for some years ^20–41^.

### Added value of this study

For all the above reasons, we proposed to experiment RICOVAI-19, a disruptive Artificial Intelligence (AI) technological support, consisting of a smartphone application questionnaires combined with a multi-parameter device which are capable of measuring certain physical and vital parameters in order to reduce access to the emergency room, if not necessary, and allow doctors to continuously monitor the health status of patients at home, prioritizing hospital evaluation / admission based on clinical severity status.

Compared to the “black box” operation of some devices that can be traced back to the concept of “automatic decisions” where the software decides autonomously, without providing explanations, the transparent operation adopted by RICOVAI-19 AI multiparametric artificial intelligence system enables the so-called “ augmented decisions”: the software does not decide autonomously, but allows a human being expert in a certain field, for example a doctor, to make a decision based on the evidence of the correlations discovered by the model and to use this additional information by integrating them with the baggage of their previous knowledge. The combinations of critical values of the different parameters are represented by tools that allow you to assign scores and view the prognostic relevance. Such example of Valuable Based Healthcare IoMT, able to orient towards clinical appropriateness, was used in RICOVAI-19 study by monitoring system using Artificial Intelligence, a “wearable” multi-parameter system that releases clinical stability indexes in the decision making process of respiratory syndromes by sending data from a smartphone application.

This system is based on methodologies that use Machine Learning or “the science that enables computers to learn, without having been explicitly programmed for this”. These are software based on mathematical algorithms that simulate inductive reasoning, learning from data and generating predictive models.

The added value is more consistent when RICOVAI-19 achieves and optimizes a network of clinical excellence being all the actors (hospitals, General Physician-GP- and patients) completely advised of any action, planning and sharing suggestions the AI system is producing. As a consequence, the quality of patient care and the doctor clinical workload will assume a virtuous meaning.

## 2. Methods

The RICOVAI-19 pilot feasibility study was classified as a prospective, open, not randomized, not pharmacological study obtaining the local Ethical Committee approval on 4^th^ March 2021 (N. 2021-30). The study was performed on the population of subjects greater than 18 years old with suspected or known COVID-19 who live in Offagna, a small town in the province of Ancona, Italy.

The RICOVAI-19 monitoring and measurement system consist of a software application accessible by a smartphone application, for the manual insertion of clinical variables and of a multi-parameter medical device, which also allows automatic insertion of the measured parameter values - CE marked “medical device” (class IIA), telemedicine platform (Class I) compliant with the GDPR – (by Aditech-Adilife srl, Italy). The primary objectives of the study were classified as follow: 1) Adherence, accessibility and satisfaction of citizen to using a dedicated AI software applicative; 2) capacity of interaction of the applicative with territory care plan and 3) capacity of interaction of the applicative with the hospital intervention plan. Among the secondary objectives we identified monitoring, using an AI applicative, the appropriateness of hospital access to the Emergency Department confirming the admission of patients with known or suspected SARS-CoV-2 (COVID-19); train the AI system.

The assessment of the objectives takes place releasing both individually (inhabitants), institutionally (hospital) and territory (GPs), a system consisting of software licenses - with access via App and web portal - equipped with a dedicated multi-parameter sensor device whose usage is facilitated by a direct connection between devices or by manual entry. The multi-parameter device that facilitates the insertion of some of the parameters necessary for the evaluation (6 parameters) can also be used, if properly configured, by other members of the family unit. The system aims to complement the standard clinical evaluation model already in place.

The system is not intended to evaluate the effectiveness of an intervention. It intends instead to provide an AI decision support algorithm according to good clinical practice. The physicians will always act according to the guidelines of good clinical practice and unconditionally from the results of the decision support AI algorithm solution (by Almawave S.p.A, Italy).

The validation of the set objectives will be given by the level of accuracy of the model. Therefore, if the validation set relative to the clinically observed values of the stability indicator is respected by the model for a proportion greater than 95%, then the accuracy is decided to have reached the desired goal. Four phases were taken into account in the study: 1) enrollment of subject by the Gp; 2) smartphone with AI applicative onboard and sensor delivery to the subject; 3) activation of subject enrolled and 4) twentyone days monitoring. Once the activation has been terminated, the subject can start the monitoring and the system alert all operators. The AI-based CSI of each patient, updated at any measure, is visible on a dashboard telemedicine platform which represent the front-end of the whole use-case patient journey (Figure 1). The RICOVAI-19 app provides recommendations on the timing and frequency of measurements, depending on the CSI value. In fact an active control room monitors patient via CSI and, based on its value, contacts GP /subject depending on type of action to be done. Hospital personnel and GP can always check the clinical condition and patient journey anytime by viewing the patient data on the dashboard applicatives in a web portal or mobile app. The control room define the end of monitoring when the “stable” CSI has been confirmed from at least three days before the 21^st^ day of participation of patient to the survey. A logistic service takes all the action to recover the AI-based applicative and sensor from the patient to allow sanification and stock.

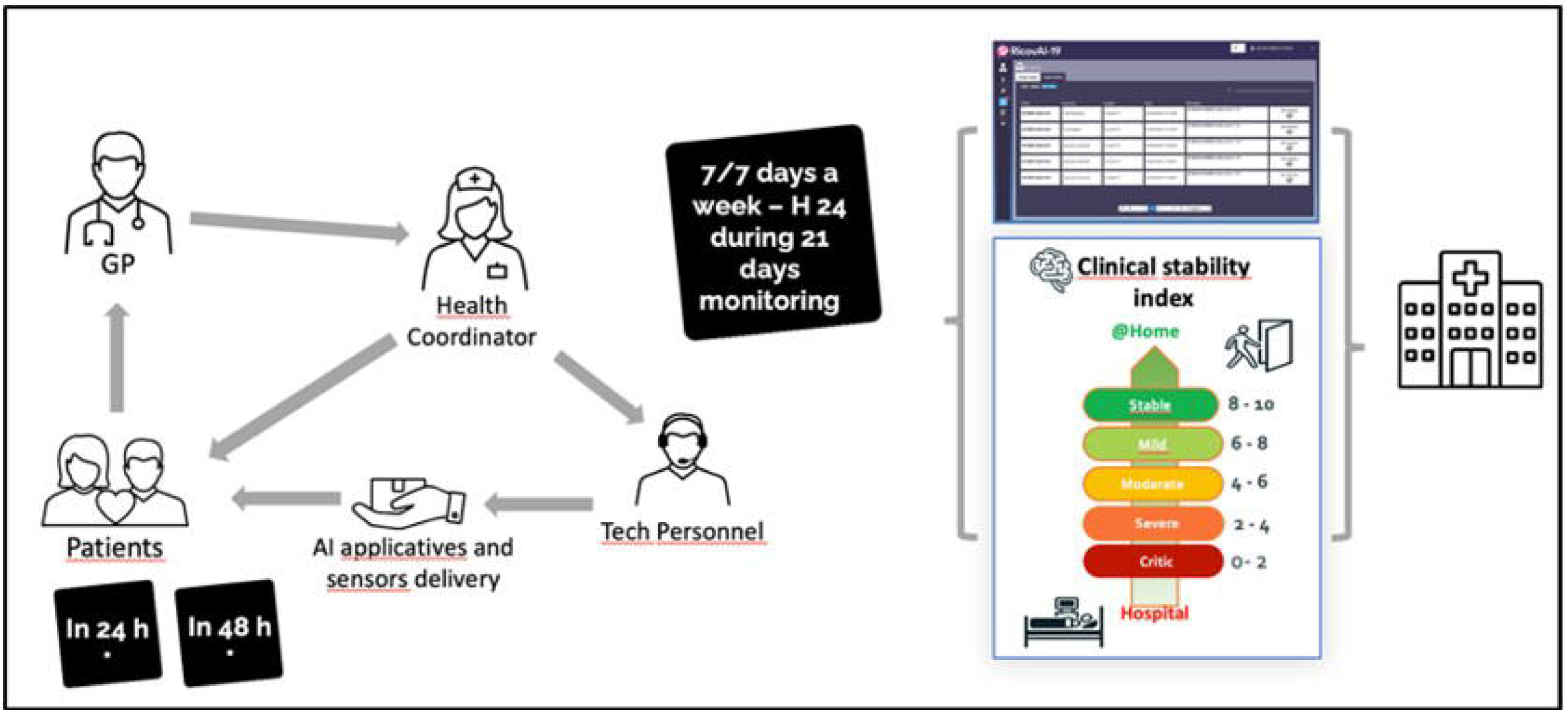

Patients engaged into this monitoring campaign (promoted by Vivisol srl, Italy) fill out a questionnaire twice a day through the smartphone application. Within the questionnaire, patients must answer a predefined set of questions regarding their health current state, in particular regarding the presence of symptoms attributable to a Covid infection and their possible severity, the pre-existence of comorbidities and risk factors, and regarding contact and exposure to any suspected or confirmed cases of Covid (see Table 1). After completing the questionnaire, the measurement of body parameters follows (see Table 2) and, on the basis of all the information collected, the application calculates and shows the patient clinical stability index (CSI), then providing the doctor the indications on subsequent behaviors that the patient must update.

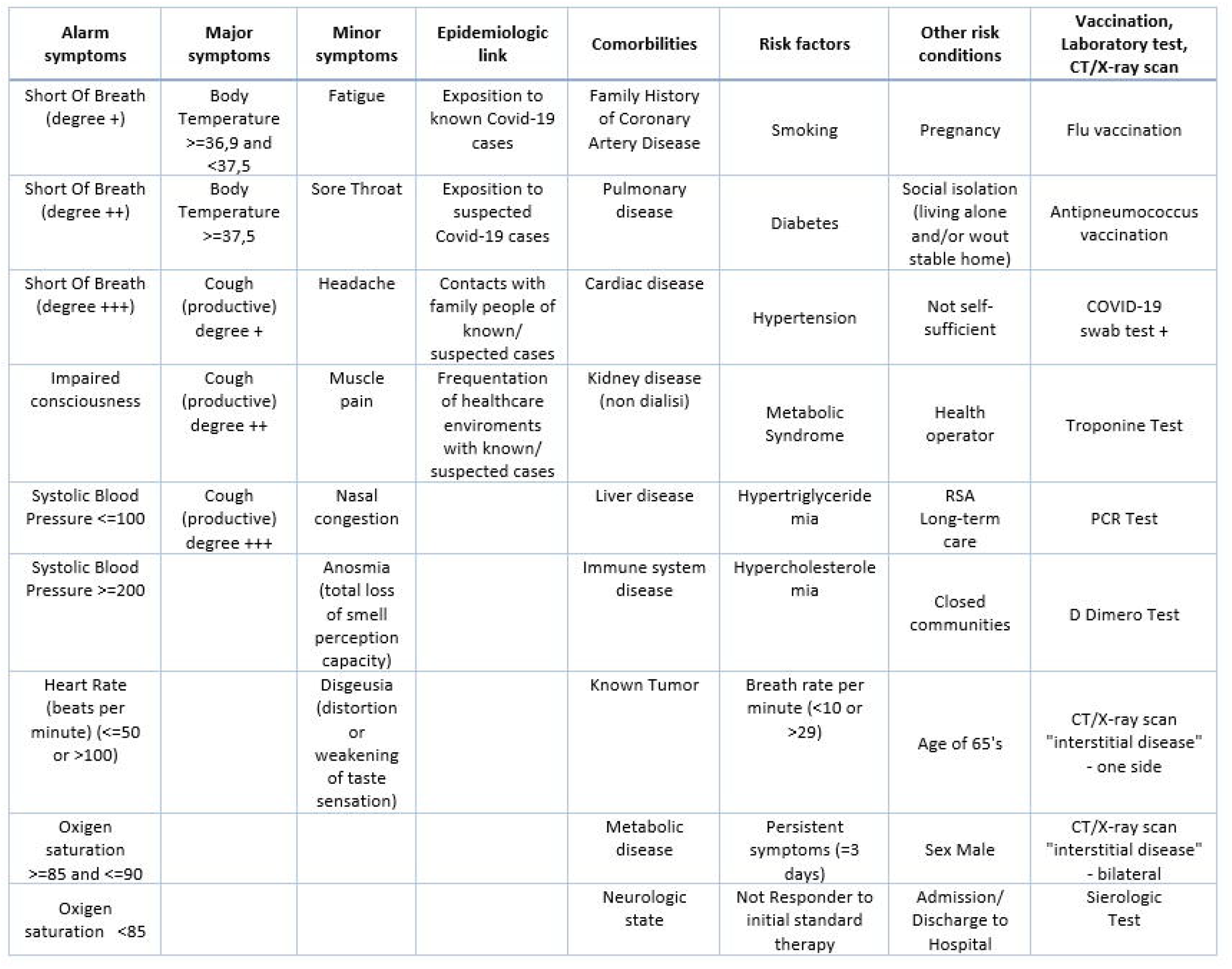

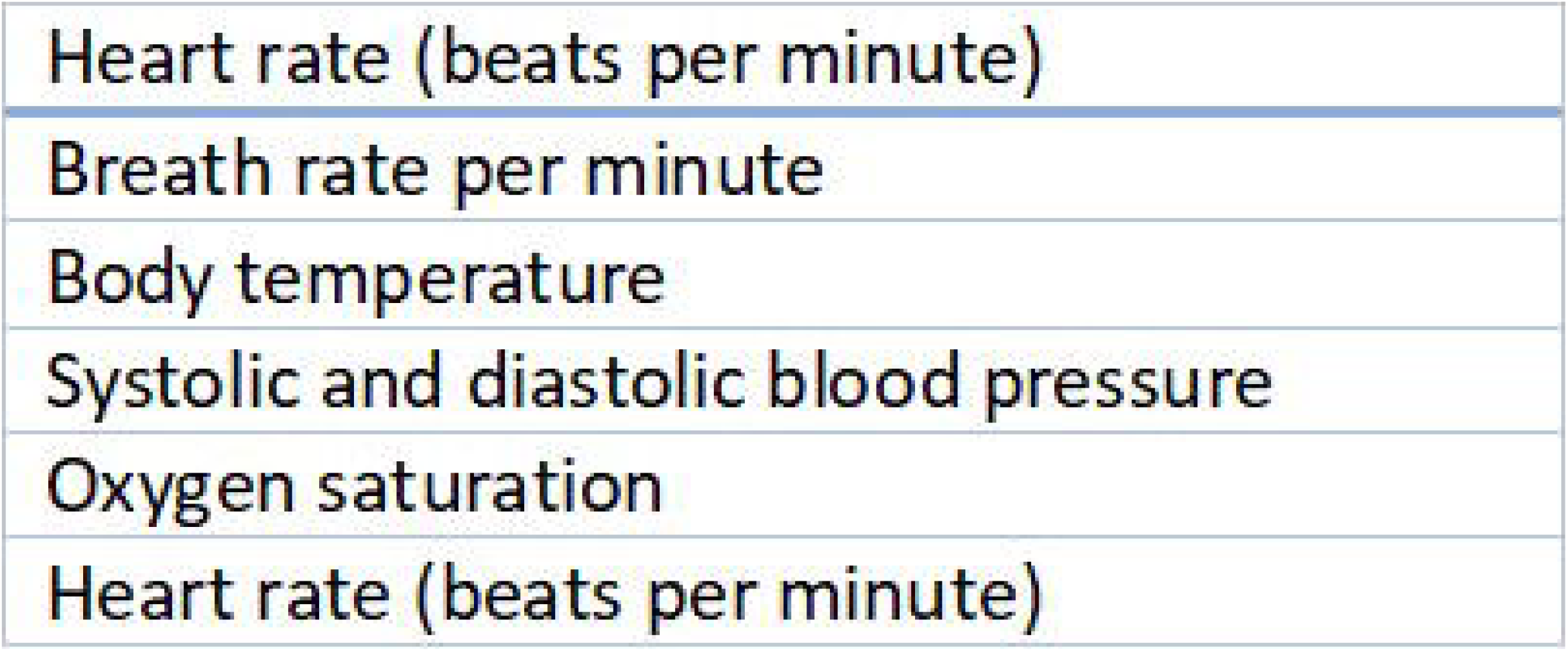

The CSI is represented by a numerical value between 0 and 10 that allows to define the patient’s state of health, with respect to any contagion from Covid. A CSI value between 0 and 2 corresponds to a critical status, while a value greater than 7 corresponds to a stable state of health of the patient. The CSI is also identifying the likelihood of negative evolution of the syndrome (see Table 3).

| Clinical Stability Index | Risk Cluster | Risk Cluster Likelihood |
| --- | --- | --- |
| 0.00-2,00 | Critical | High likelihood of negative evolution of the syndrome – Definitely high risk |
| 2,01-3,25 | Severely Unstable | Moderate-severe likelihood of negative evolution of the syndrome – Severely increased risk |
| 3,26-4,50 | Moderately Unstable | Intermediate-moderate likelihood of negative evolution of the syndrome – Moderately increased risk |
| 4,51-7,00 | Mildly Unstable | Mild likelihood of negative evolution of the syndrome – Mildly increased risk |
| 7,01-10,00 | Stable | Low likelihood of negative evolution of the syndrome – Lowly increased risk |

CSI allows the doctor to monitor the patient state of health and to evaluate any worsening or improvement of the related clinical conditions.

CSI was defined using AI and specifically supervised learning techniques derived by machine learning (ML). In general, at the base of ML there is the concept of applying mathematical-computational rules to learn directly from data. The idea was to be able to study a sample of data to understand which characteristics link the independent variables to the dependent ones of the problem and to what extent. In this case, a technique called linear discriminant analysis was used ^42^. The goal of the discriminant analysis is to assign a subject to one of the homogeneous groups already identified, minimizing the risk of incorrect classification^43^.

Therefore, discriminant analysis has the same objective as group analysis, that is, to classify patients, with the difference that in this case the groups are already known ^44^.

The discriminant analysis built on real cases divides the same based on medical experience (supervised) into 5 homogeneous groups (5 levels of clinical diagnosis) Applying Fisher’s linear discriminating functions ^45^. During the application phase, the discriminant analysis allows you to define the probabilities of belonging to each group and then summarizing these probabilities in a synthetic score, named “CSI”.

### Statistical analysis

The CSI has been constructed from a sample of individuals observed at three distinct moments in order to build the retrospective time series. About 1,500 observations were obtained for which 58 independent variables were collected and valuated (see table 1). Based on the medical evaluation, that each of those observations has assigned a value to the CSI. The observations were classified according to the severity of the stability indicator in 5 homogeneous groups, the Clinical Stability Index Risk Clusters (see table 3).

The predictive algorithm of the CSI has been identified through supervised learning applying the linear discriminant analysis.

Reclassification of the observations used for the estimation of linear discriminant functions showed an excellent level of reassignment to the starting groups (Table 4).

| Arrival group | 1 | 2 | 3 | 4 | 5 |  |
| --- | --- | --- | --- | --- | --- | --- |
| Starting group | Stable | Mild | Moderate | Severe | Critic | Total |
| 1 – Stable | 30 |  |  |  |  | 30 |
| 2 – Mild | 5 | 90 |  |  |  | 95 |
| 3 – Moderate |  | 10 | 100 | 15 |  | 125 |
| 4 – Severe |  |  | 10 | 180 | 5 | 195 |
| 5 – Critic |  |  | 5 | 50 | 1.000 | 1.055 |
| Total | 35 | 100 | 115 | 245 | 1.005 | 1.500 |

| Error Count Estimates |  |  |  |  |  |  |
| --- | --- | --- | --- | --- | --- | --- |
| Groups | 1 | 2 | 3 | 4 | 5 | Total |
|  | Stable | Mild | Moderate | Severe | Critic |  |
| Badger | 0,0000 | 0,0526 | 0,2000 | 0,0769 | 0,0521 | 0,0763 |

In summary, the reassignment error is very low (7.6%) and is limited to contiguous clusters in terms of risk. It should be noted that of the 1,500 observations used, 1,200 observations (80% of the population) were used in the construction phase of the discriminating functions, while 300 observations (20% of the population) were used as a test sample of the results.

In this way, the possibility of a test on a significant sample of real cases not used in the construction phase of the methodology was guaranteed.

In the application phase, the probabilities of belonging to the 5 homogeneous groups are calculated according to the following formula:

where:

i is the cluster number

X is the vector of discriminating variables

*C*_*i*_ is the vector of the coefficients of the i-th linear discriminating function.

Based on the probabilities of reassignment to homogeneous groups, the Clinical Stability Index is calculated by applying the following formula:

In order to identify the behavioral patterns that are as close as possible to individuals and representative of the various specificities that may occur, trainings were carried out with discriminating analysis on increasingly numerous samples of observations.

The aim is to specialize machine learning models. For such training, accuracy will be evaluated until the desired result is achieved.

In the clinical investigation phase, the RICOVAI-19 application was used 7. 386 times from the 129 patients of Offagna. The accuracy of the AI model was carried out by the doctors involved in the clinical trial, who reported only 9 cases that were not caught by discriminating functions.

Thanks to the medical evaluation that for each of these observations has reassigned the relative “expected” value of the Clinical Stability Index, it was possible to replicate - through supervised learning - the predictive algorithm of the same indicator.

In a sub-analysis, RICOVAI-19 demonstrated to be able to allow and to analyze the behavioral models of the subjects subjected to monitoring regarding the behaviors of the monitored subjects.

In this analysis, **decision trees** has been used. When we talk about the decision tree we refer to a powerful and widespread **predictive machine learning** technique that is used to predict variables that can be classified as discrete (and then we talk about classification) or continuous (in this case regression). A popular algorithm that allows us to achieve this goal is definitely the **CART** (Classification And Regression Trees) ^46^Applying the decision trees (see figure 2), the behaviors of the subjects monitored by RICOVAI-19 emerged:

- Subjects with incorrect measurements tend to quickly repeat the detection, after reading a low Clinical Stability Indicator;
- A subject monitored at home, worried by persistent symptoms for more than three days, carries out the new detection within 6 hours, while the other subjects at home tend to follow the indications of the app and carry out the 2 daily surveys.

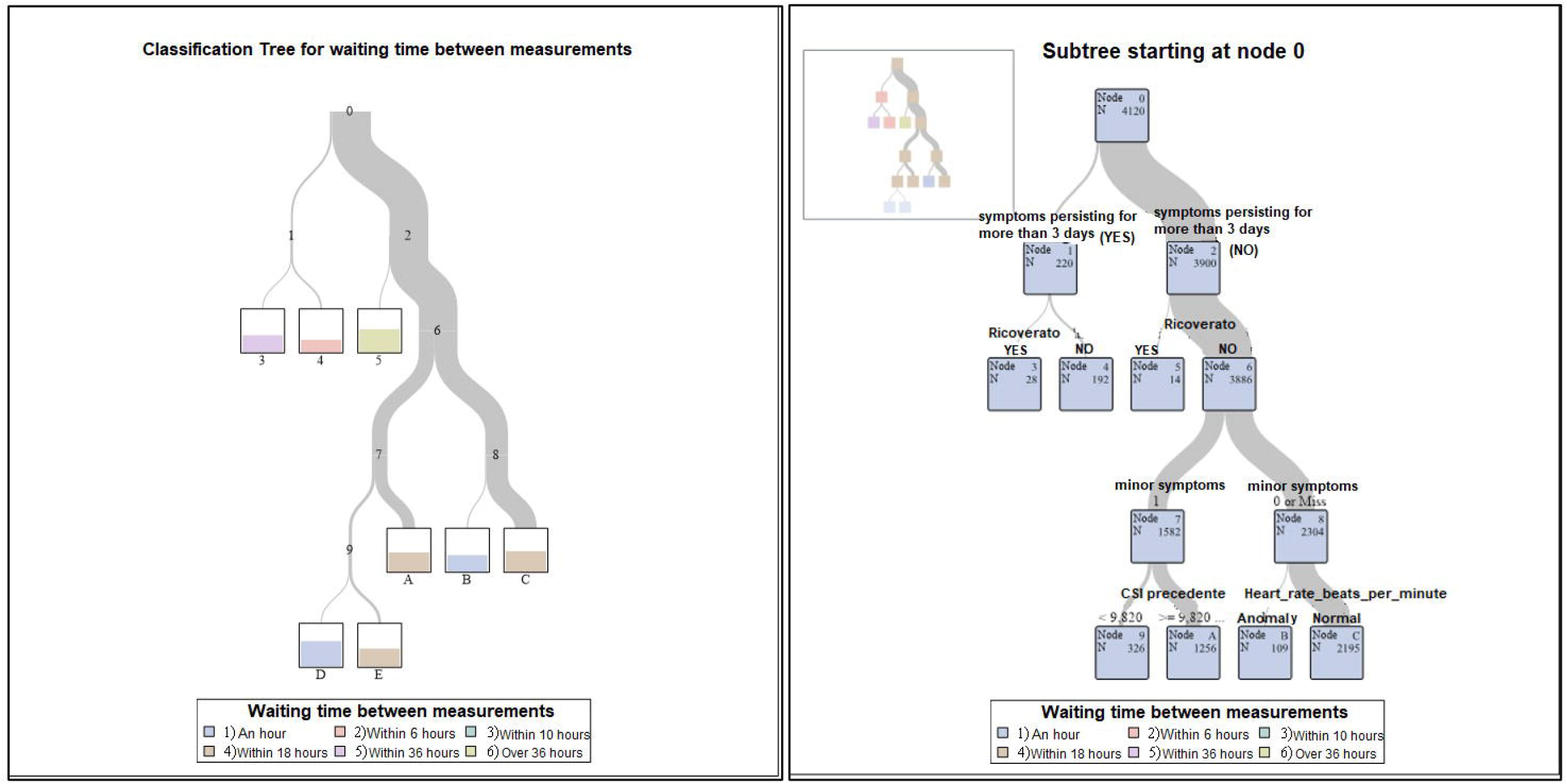

## 2. Results

From March 22, 2021 to October 22, 2021 we evaluated 129 subjects, 60 males (mean age 59 ± 14.5 years) and 69 females (mean age 53.6 ± 15.5 years). All subjects enrolled into the study were monitored for 21 days as shown in monthly recruitment (Figure 3).

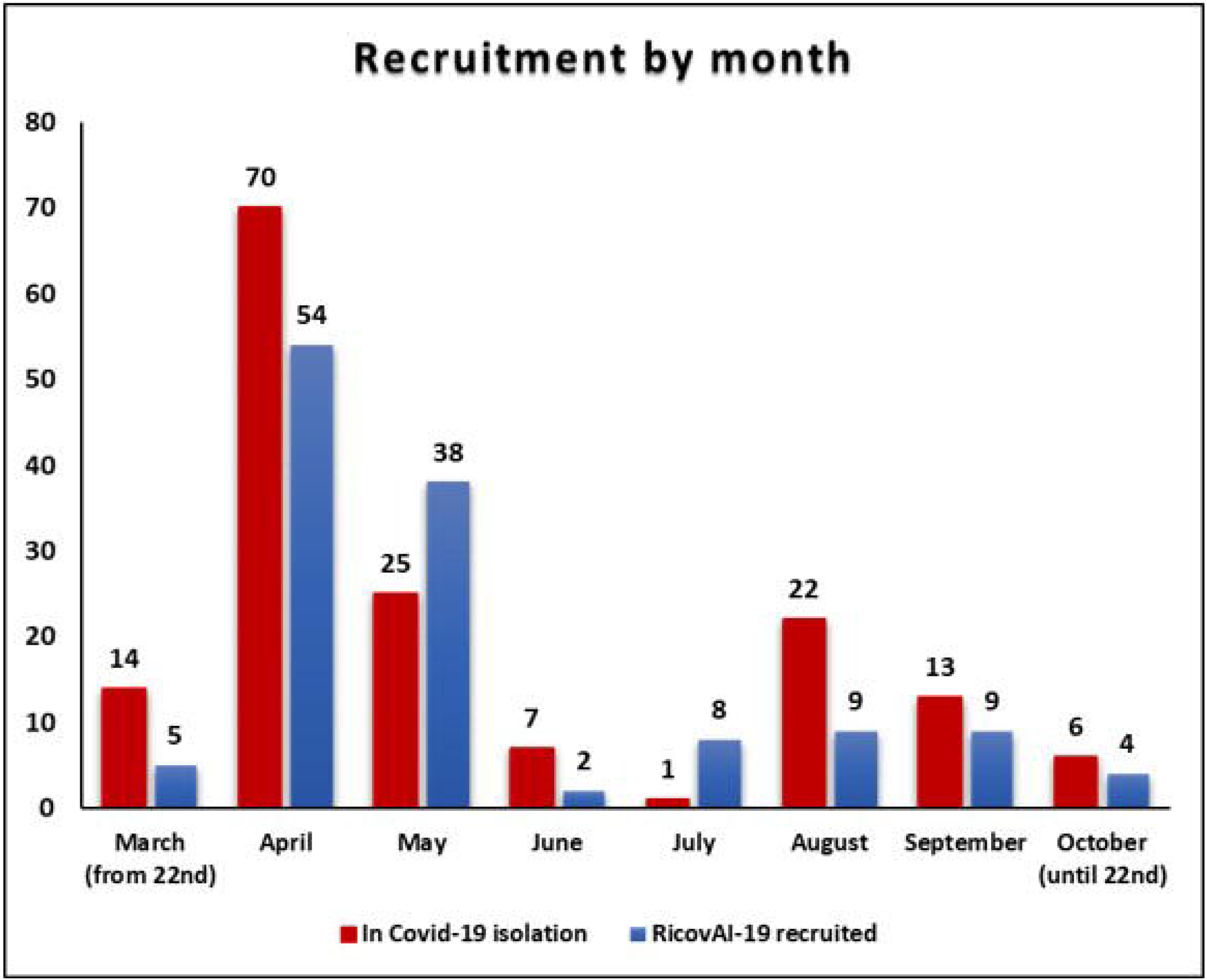

Among the objectives of the study the adherence to AI-based digital applicative was very good being > around 40%. This means that more than 40% of targeted population were enrolled into the study once contacted by GP. In fact, during the 6-months period of clinical investigation in the Municipality of Offagna, 38 subjects were positive at swab test and remained isolated while 120 subjects resulted as strictly case contact to a positive person and then remained in quarantine. Of the 158 recruits, 129 subjects (equal to 82%) have joined and participated in the experimentation of the RICOVAI-19 system, a percentage much higher than the target of 20-40%.

Regarding the accessibility and usability of digital app, the mean value was reported to be > 60%. 71% of those enrolled carried out a number of daily measurements equal to or greater than those suggested by the application (Figure 4).

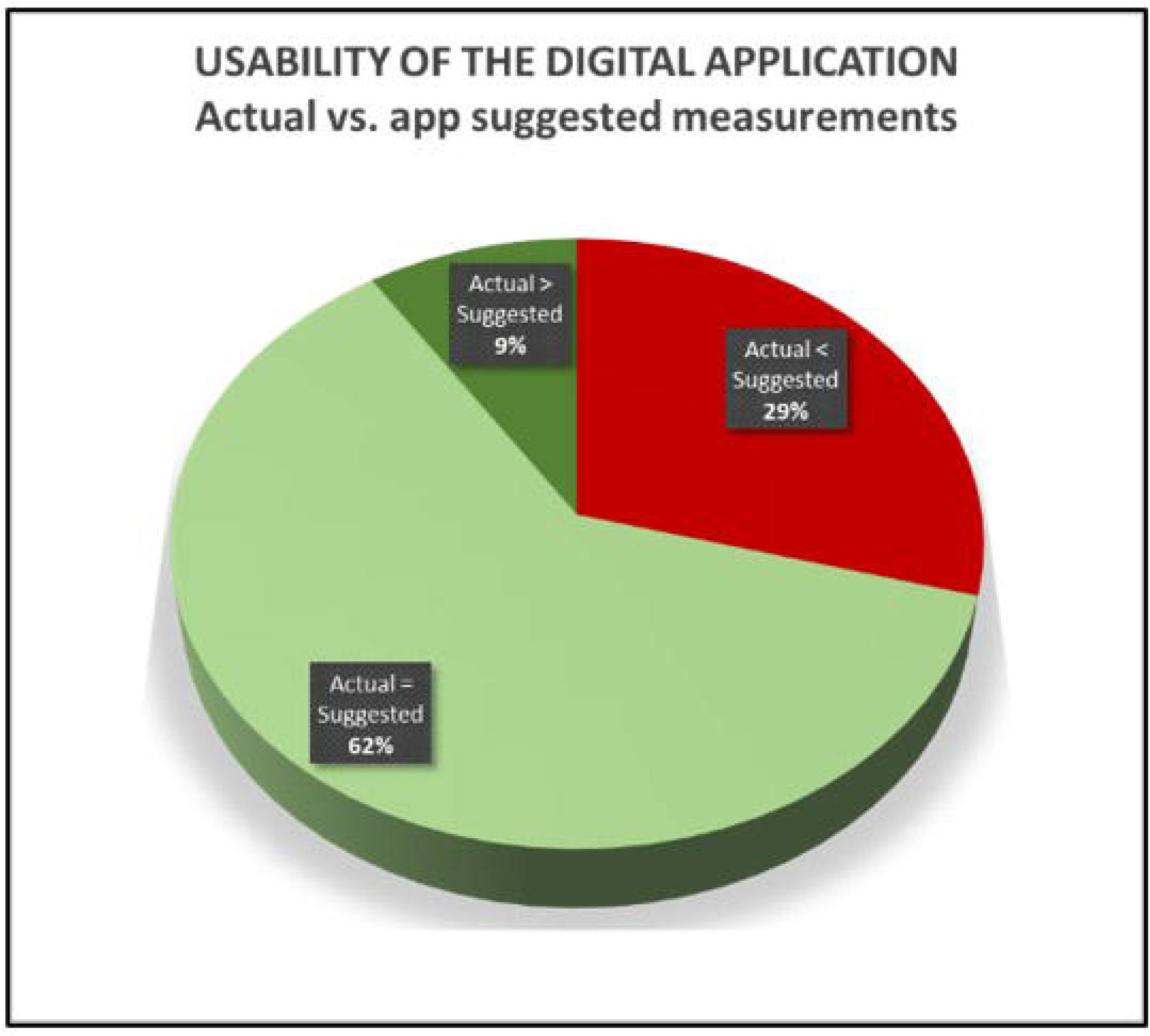

The AI model continuous training experienced CSI values > 95% of patients enrolled.

The total of patients recruited (129) carried out 7,386 surveys, for an average of 57 surveys x recruited subject in the whole period of monitoring (an average of 3 surveys per day). Of them only 9 cases (0.1%) had an expected CSI different from the RICOVAI-19 result.

In addition, the training phase led to the identification and insertion of 7 new information in the RICOVAI-19 questionnaires (of which 3 active in the discriminant analysis for the calculation of the CSI).

To have statistical consistency within each CSI cluster we assumed that at least 200 observations were needed be acquired. Figure 5 demonstrates that aim has been successfully reached except in the “CSI 2-4; severely unstable” class (173/200).

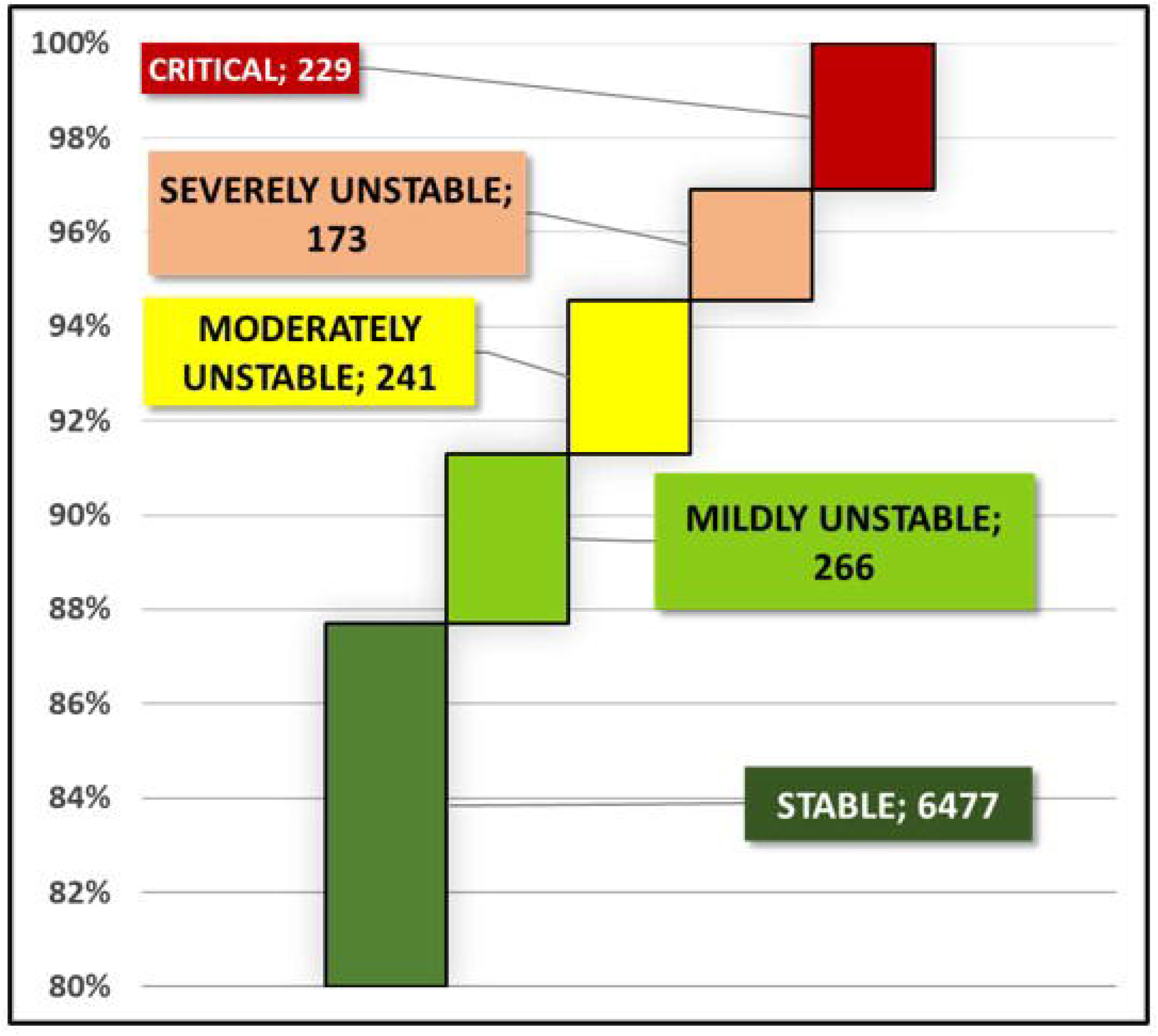

The interaction patient-AI-based digital applicative with GPs successfully resulted greater than 3 per patient (Table 5). The interaction of the digital application with the treatment plan has been defined as “every time the system provides clinical information and vital parameters (electronically) to GP.

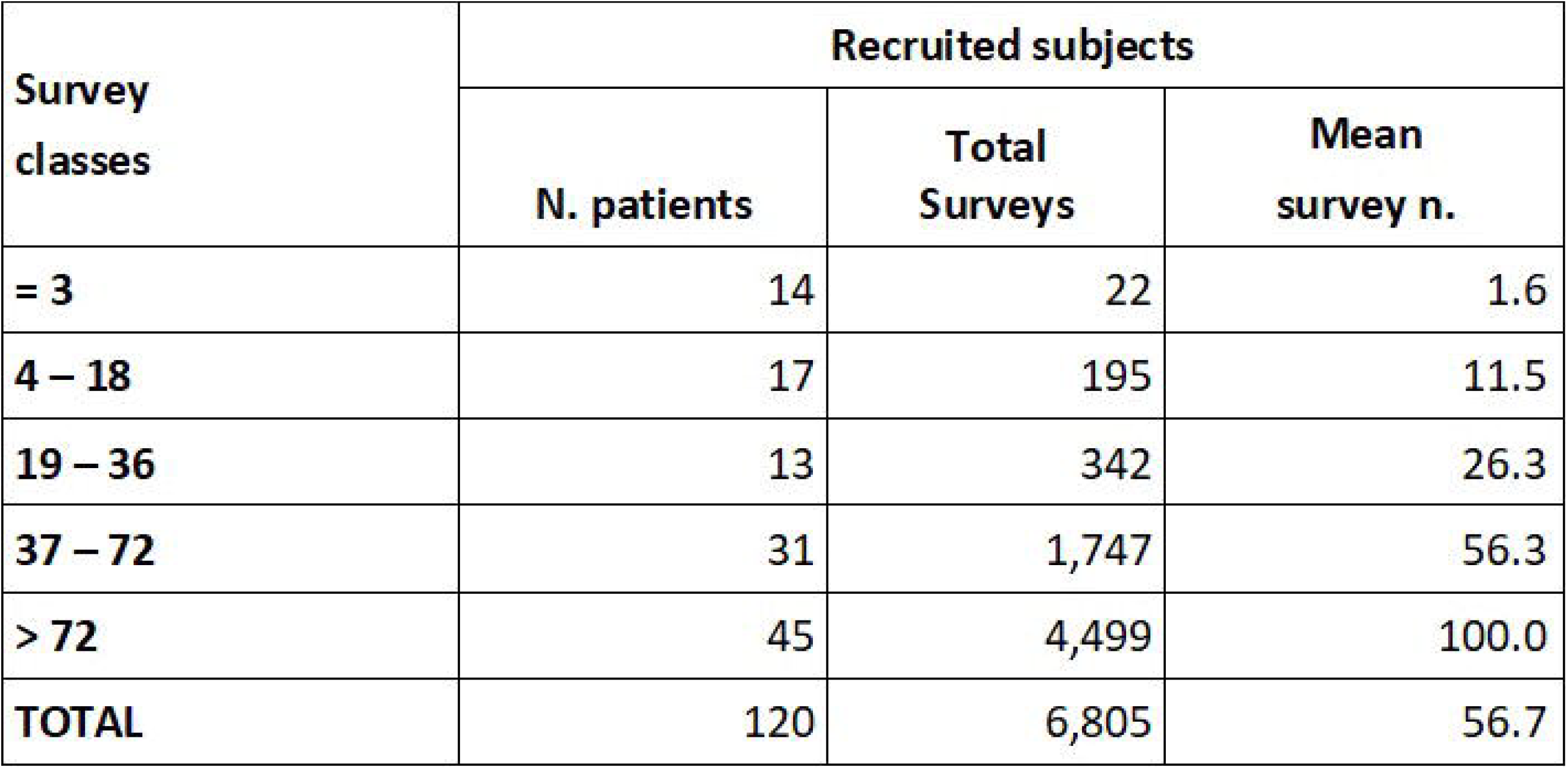

By the Hospital perspective, the interaction resulted of a significant impact being greater than 3 surveys per subject (Table 6). Differently from previous use-case, here the interaction of the digital application with the treatment plan is defined as “every time the system provides clinical information and vital parameters (electronically) to in-hospital Specialist”.

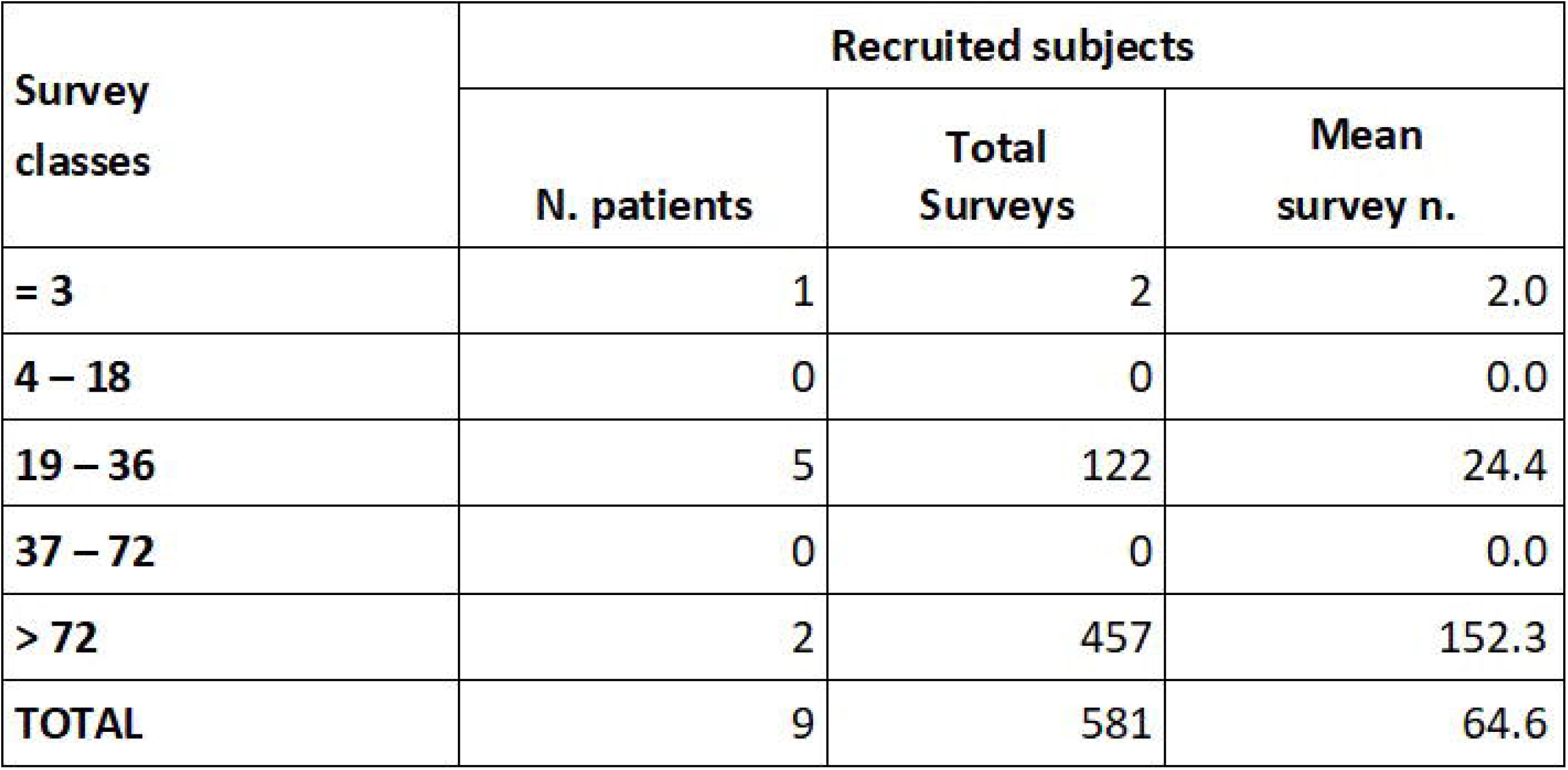

CSI was reported of high efficacy in the whole care and monitoring patient journey, from activation to the treatment of a possible acute event. Only 1 patient out of 128 accessed the emergency department (ED) on the recommendation of the family doctor based on good clinical practice but not derived from AI-based CSI result (7.3). In fact the patient was discharged with negative swab test from ED. As a consequence we have hypothesized to state that when high CSI is observed, between 7 and 10 (stable patient), no indication to send patient to the ED at all. The rest of 128 subjects were monitored at home and none of them were admitted to the Hospital.

## 3. Discussion

The AI algorithm used is patient-centered tool. The term “patient-centered” implies a stronger and more active patient. Since the beginning of machine learning continuous training of the algorithm, this healthcare organization model was reinforced and filtered by doctors interaction and it was made possible also by patients and their families contribution to make the system informed having then all shared decisions on line with subjets expectations at the end. We prefer to name “smart patients” all those subjects who adhere to the study being at the centre into a pro-active engagement.

People become more aware of the factors that affect their health and each person will progressively become more responsible for their management. We therefore refer to a smart patient, who is a committed, proactive and questioning patient, who would like to collaborate with his healthcare team to make good <u>decisions</u> about diseases and treatments, to seek relevant information, to be informed and educated about his Health problems. Smart patient condition is monitoring by visits, being able to provide healthcare professionals with a more complete description of their disease or condition. To this end, they collect personal health-related data using wearable devices and network technologies.

RICOVAI-19 guarantees mobility and continuity: patients are able to access and / or collect information and data wherever they are, at any time.

The citizen does not have a homogeneous “participatory behaviour”. There are subjects who adhere to the protocol with repeated surveys several times per day and others who instead perform only a few and discontinuous surveys throughout the observation period. The RICOVAI-19 app provides recommendations on the timing and frequency of measurements, depending on the CSI value. In this study the behaviour of the citizens involved in the RICOVAI-19 project was monitored.

The CSI ranges from 1 to 10 and is very intuitive for the patient because it is very similar to the grade on any report card. When a measure error event happens, the patient detects a significant reduction in the CSI and, as a reaction, he repeats the measurement more carefully obtaining the correct value.

AI-based CSI demonstrated to be a unique, single, shared synthetic tool able to advise prognostic information predicting a trend of the stable condition over time. This approach explains how important patient engagement and empowerment are in a sort of participative medicine.

RICOVAI-19 model allows doctors to monitor patients in real time, to define personalized therapies and to avoid overcrowding in the ED and hospitalization.

AI-based ehealth clinical decision support, by sharing the uniform data and update in real time, give a meaning of a virtuous continuous care model as the desirable collaborative integration between territory healthcare and referral hospital during pandemic.

## Data Availability

All data produced in the present work are contained in the manuscript

